# Educational level as a cause of type 2 diabetes mellitus: Caution from triangulation of observational and genetic evidence

**DOI:** 10.1101/2021.07.17.21260688

**Authors:** Nat Na-Ek, Juthamanee Srithong, Authakorn Aonkhum, Suthida Boonsom, Pimphen Chareon, Panayotes Demakakos

**Author notes:** **Corresponding author** Dr Nat Na-Ek, Department of Pharmaceutical Care, School of Pharmaceutical Sciences, University of Phayao, Phayao, Thailand.

## Abstract

**Background:** Education might be causal to type 2 diabetes mellitus (T2DM). We triangulated cohort and genetic evidence to consolidate the causality between education and T2DM.

**Methods:** We obtained observational evidence from the English Longitudinal Study of Ageing (ELSA). Self-reporting educational attainment was categorised as high (post-secondary and higher), middle (secondary), and low (below secondary or no academic qualifications) in 6,787 community-dwelling individuals aged ≥50 years without diabetes at ELSA wave 2, who were followed until wave 8 for the first diabetes diagnosis. Additionally, we performed two-sample Mendelian randomisation (MR) using an inverse-variance weighted (IVW), MR-Egger, weighted median (WM), and weighted mode-based estimate (WMBE) method. Steiger filtering was further applied to exclude single-nucleotide polymorphisms (SNPs) that were correlated with an outcome (T2DM) stronger than exposure (education attainment).

**Results:** We observed 598 new diabetes cases after 10.4 years of follow-up. The adjusted hazard ratios (95%CI) of T2DM were 1.20 (0.97-1.49) and 1.58 (1.28-1.96) in the middle- and low-education groups, respectively, compared to the high-education group. Low education was also associated with increased glycated haemoglobin levels. Psychosocial resources, occupation, and health behaviours fully explained these inverse associations. In the MR analysis of 210 SNPs (R^2^=0.0161), the odds ratio of having T2DM per standard deviation-decreasing years (4.2 years) of schooling was 1.33 (1.01-1.75; IVW), 1.23 (0.37-4.17; MR-Egger), 1.56 (1.09-2.27; WM), and 2.94 (0.98-9.09; WMBE). However, applying Steiger filtering attenuated most MR results toward the null.

**Conclusions:** Our inconsistent findings between cohort and genetic evidence did not support the causality between education and T2DM.

**Key messages:** *What is already known on this subject?:*

- Several pieces of evidence suggested that education attainment might play a causal role in the occurrence of T2DM.

*What does this study add?:*

- Our observational evidence suggested no direct impact of education on the risk of T2DM. The observed inverse associations were mediated through insufficient psychosocial resources, low occupation class, and unhealthy behaviours due to low education.
- In contrast, the genetic evidence suggested no causal association between education and the risk of T2DM. Notably, the significant associations from our genetic evidence resulted from the invalid genetic instrument used in the analysis.
- The observational and genetic evidence was inconsistent; therefore, our triangulated evidence did not support a causal role of education in the occurrence of T2DM.

## Introduction

Type 2 diabetes mellitus (T2DM) is a significant global burden affecting more than 422 million people worldwide, and its prevalence will reach 7,079 per 100,000 by 2030.[1] In some countries, the increased prevalence reflects a better survival rate, while the incidence of T2DM is still rising in others.[2]

Currently, T2DM is incurable. Thus, prevention is crucial. An effort has been put into clinical risk modification, such as weight reduction and smoking cessation.[3] However, it has been suggested that these clinical factors attributed to only one-third of total diabetes risks.[4] Therefore, the residual factors are worth further investigation.

Previous observational studies have shown an inverse association between educational level and the risk of T2DM.[5–8] However, residual confounders and reverse causality limited the establishment of causality. Moreover, scarce evidence had examined the relationship between education and glycated haemoglobin (HbA1c) levels,[9] which are the biomarker of prediabetes and well-established cardiovascular risk.[10] Furthermore, findings from genetic (Mendelian randomisation) studies are equivocal.[11–14]

This study aims to investigate the causal effect of education on the risk of T2DM and HbA1c levels by comparing results from two different study designs – an approach called ‘triangulation of evidence’.[15] Triangulated findings may complement the limitations of each other and provide a more solid conclusion. The two methods being used here are cohort study and Mendelian randomisation (MR). In brief, MR uses single-nucleotide polymorphisms (SNPs) as a proxy of exposure. This genetic proxy is less likely to be associated with confounders due to its random allocation according to Mendel’s law of independent assortment.[16–18] Additionally, we also examine the causal pathway linking educational level with the risk of T2DM and HbA1c levels.

## Methods

This report followed the STrengthening the Reporting of OBservational Studies in Epidemiology (STROBE) guidance of cohort studies and its extension to Mendelian randomisation (STROBE-MR) (Table S1-S2).[19]

### Cohort evidence

#### Data source and study population

We used the English Longitudinal Study of Ageing (ELSA) data: a prospective cohort study of nationally representative community-dwelling individuals aged ≥50 years. At ELSA wave 1 (2002-03), samples included all consenting people who participated in the Health Survey for England (HSE) in 1998, 1999 and 2001. Subsequent follow-up interviews and health examinations take place regularly at two- and four-year intervals, respectively. More information on ELSA can be found at http://www.elsa-project.ac.uk/.[20]

We used data from ELSA wave 2 (2004–05), which comprised follow-up interviews and health examinations and constituted the baseline of our cohort study. Of 11,391 participants in ELSA wave 1, 8,780 participated in ELSA wave 2, of whom 7,666 consented to the health examination. Our final analysis included 6,786 individuals without a history of diabetes in ELSA wave 2. To make the results comparable to genetic evidence, only white participants (97.7% of core samples in ELSA wave 2) were included in the analyses (Figure S1).

#### Educational level

Educational level was the self-reported highest educational qualification obtained by ELSA wave 2, further classified into three groups as implemented by a previous ELSA study.[5] A high educational level was defined as a university degree, other higher or post-secondary education, and A-level education (n=2,218), whereas a middle educational level included a Certificate of Secondary Education (CSE) and similar foreign qualifications (n=2,106). Individuals with below secondary education or without educational qualifications were grouped as low educational level (n=2,462).

#### Type 2 diabetes mellitus (T2DM) and glycated haemoglobin (HbA1c) levels

The primary outcome was the self-report physician-diagnosed diabetes up to ELSA wave 8 (2016/17). To minimise misclassification bias, we included participants with HbA1c levels ≥6.5% at least twice in a diabetic group as suggested clinically.[10] The secondary outcome was the trajectory of HbA1c levels measured at ELSA wave 2, 4, 6, and 8. Notably, HbA1c measured in ELSA before October 2011 was calibrated using Diabetes Control and Complications Trial (DCCT) standards, replaced by the International Federation of Clinical Chemistry (IFCC) standardisation afterwards. Details of quality control of HbA1c measured in ELSA has been published elsewhere.[21]

#### Covariates

We collected all covariates at baseline, mostly self-reported, except for body mass index (BMI). These covariates included age (years), age^2^, sex (i.e., male and female), marital status (i.e., single, married, and separated divorced or widowed), depressive symptom (i.e., Center for Epidemiologic Studies Depression [CESD] score ≥4), occupational class (i.e., managerial or professional, intermediate, and routine or manual occupation), BMI (i.e., normal, overweight, and obese), smoking status (i.e., never, ex-, and current smoker), alcohol drinking (i.e., never or almost never, 1-2 times a month, 1-2 times a week, and daily or almost daily). Moreover, childhood socioeconomic position (SEP) was obtained and categorised into four groups according to father’s main job when participants aged 14 years: high (i.e., managerial-, professional-, administrative occupations, or business owners); middle (i.e., trade- or services related occupations); low (manual or casual occupations, unemployed, sick and disabled); and miscellaneous (i.e., armed forces and retired). According to directed acyclic graphs (DAGs) adapted from Hamad *et al*.[22] and Liang *et al*.[14] only age, sex, and childhood SEP were considered confounders, whereas the rest were mediators (Figure S2).

#### Statistical analysis

Sample baseline characteristics were explored according to educational groups using descriptive and inferential statistics, as appropriate. We created a Kaplan-Meier plot for the cumulative incidence of T2DM of each group and compared it by log-rank test.

The association between educational levels and the risk of T2DM was examined using a Cox-proportional hazards model with high education as a reference group. To investigate potential causal pathways, models were adjusted for each set of covariates as follows: confounding factors (i.e., age, sex, and childhood SEP); psychosocial resources (i.e., depressive symptom and marital status); occupational class (i.e., occupation); health behaviours (i.e., BMI, smoking, alcohol drinking, and physical activity); and a final model that was accounted for all covariates. The proportional hazards assumption was checked by Schoenfeld residual statistic and log-minus-log plots. The multicollinearity of covariates was examined by calculating variance inflation factor (VIF). Covariates with missing data (mostly missed <5%, Table 1) were multiply imputed by chain equation (MICE) with 50 imputations.

**Table 1.** Baseline characteristics of included participants

| Characteristics | Educational levels |  |  | Total |
| --- | --- | --- | --- | --- |
|  | High | Middle | Low |  |
| N | 2,218 | 2,106 | 2,462 | 6,786 |
| Male | 1,220 (55.0) | 851 (40.4) | 890 (36.2) | 2,961 (43.6) |
| Age (years) | 63.6 ± 8.8 | 65.2 ± 9.2 | 69.6 ± 10.1 | 66.3 ± 9.8 |
| High occupational class | 1,400 (63.1) | 503 (23.9) | 240 (9.8) | 2,143 (31.6) |
| Missing | 11 (0.5) | 16 (0.8) | 65 (2.6) | 92 (1.4) |
| High childhood SEP | 1,123 (46.3) | 745 (30.9) | 468 (15.6) | 2,336 (29.8) |
| Missing | 96 (4.0) | 96 (4.0) | 126 (4.2) | 318 (4.1) |
| Non-married (separated, divorced, widowed) | 464 (20.9) | 583 (27.7) | 904 (36.7) | 1,951 (28.8) |
| Elevated depressive symptoms | 213 (9.6) | 283 (13.4) | 457 (18.6) | 953 (14.0) |
| Missing | 10 (0.5) | 10 (0.5) | 24 (1.0) | 44 (0.7) |
| Never smoked | 874 (39.4) | 784 (37.2) | 852 (34.6) | 2,510 (37.0) |
| Missing | 10 (0.5) | 5 (0.2) | 13 (0.5) | 28 (0.4) |
| Never/ almost never drunk | 352 (15.9) | 507 (24.1) | 948 (38.5) | 1,807 (26.6) |
| Missing | 10 (0.5) | 5 (0.2) | 15 (0.6) | 30 (0.4) |
| High physical activity level | 596 (26.9) | 423 (20.1) | 345 (14.0) | 1,364 (20.1) |
| Missing | 10 (0.5) | 12 (0.6) | 25 (1.0) | 47 (0.7) |
| Body mass index (kg/m<sup>2</sup>) | 27.3 ± 4.6 | 27.5 ± 4.7 | 28.1 ± 4.9 | 27.7 ± 4.7 |
| Missing | 451 (20.3) | 427 (20.3) | 699 (28.4) | 1,577 (23.2) |
| Having a history of CVDs | 1,017 (45.9) | 1,075 (51.0) | 1,408 (57.2) | 3,500 (51.6) |

**Notes** All P-values (not including a missing group) were < 0.001. Figures represent frequency (%) or mean ± SD. Abbreviations: CVDs; Cardiovascular diseases, SEP; Socioeconomic position

Additionally, we performed the following sensitivity analyses: First, we analysed only complete-case samples; second, we calculated a Bonferroni adjusted (97.5%) confidence interval to account for multiplicity. Moreover, we performed subgroup analyses according to sex, age groups (i.e., <75 and ≥75 years old), BMI groups, and smoking status.

To examine the association between education and the trajectory of HbA1c levels, we used a multilevel linear (growth curve) model, allowing for random intercepts and random slopes with unstructured covariance. The adjustment was similar to the T2DM outcome but based on a complete-case approach. The model’s validity was checked from the distribution of intercepts and slopes. Sensitivity analysis was performed by excluding participants with reporting diabetes during follow-up since they might receive antidiabetic agents that can modify HbA1c levels and distort the actual effect of education on HbA1c.

### Genetic evidence

#### Data source

All SNPs used in our study were derived from an MR-based platform as summary-level data publicly available from https://www.mrbase.org/.[23] Specific ethical approval and consents were already obtained in the original studies. Details of each genetic consortia can be found in supplementary appendices (Table S6).

#### Selection of instrumental variants

We obtained SNPs associated with years of schooling from the Social Science Genetic Association Consortium (SSGAC).[24] SNPs that reached genome-wide-significance (i.e., P-value<5*10^−8^) were selected and further pruned using linkage disequilibrium (LD)-r^2^<0.001 within a 10,000 kb window. The measuring unit of education in SSGAC was per standard deviation (SD) increase in years of schooling (4.2 years).

#### Outcomes and variants harmonisation

T2DM and HbA1c variants were taken from the DIAbetes Genetics Replication And Meta-analysis (DIAGRAM) consortium[25] and the UK Household Longitudinal Study (UKHLS),[26] respectively. Variants from different consortia were harmonised, allowing for both palindromic SNPs (i.e., a minor allele frequency [MAF] threshold of 0.3) and proxy SNPs (i.e., LD r^2^>0.8 within 10,000 kb window; Figure S6).

#### Statistical analysis

The main analysis was performed using a multiplicative random-effect inverse-variance weighted (IVW) method. For sensitivity analyses, we used the MR-Egger approach to examine and account for unbalanced horizontal pleiotropy, if any.[27] Also, we performed a weighted median and weighted mode MR. The former allowed for the invalidity of half of SNPs[28], whereas the latter minimised the false-positive rates of findings.[29] These three additional analyses were performed according to the guidelines for conducting MR investigations.[30]

Moreover, during a preliminary analysis, we observed that some of our selected SNPs had a stronger association with the outcome than exposure. Therefore, we further applied MR Steiger filtering to remove those SNPs and performed analyses accordingly.[31] To ensure the validity of the genetic instruments and processes used in our MR analyses, we also examined the association between education levels and the risk of Alzheimer’s disease (AD: obtained from the International Genomics of Alzheimer’s Project [IGAP] consortium[32]) as a positive control. This is because evidence suggested that higher education is causally related to a decreased risk of AD.[33,34]

The power of derived effect size was estimated using the method given by Hermani *et al*.[23] and Deng *et al*.[35] All analyses were performed using R version 3.6 and STATA version 16.1MP (StataCorp, LLC) with a two-sided alpha error of 5%. Since we considered T2DM and HbA1c clinically correlated, we did not adjust for multiple testing in the MR analyses.[36]

#### A conceptual framework of using the triangulation approach in this study

- If findings from the cohort study show a significant association after adjusting for the main confounders, then the true association is likely. Explanatory pathways will be further elucidated to provide insight regarding a direct path between exposure and outcome.
- The MR study is implemented to explore whether the observed association is due to causation according to our conceptual framework of MR (supplementary appendices). When evidence of causation is shown and the direction of the associations between the cohort and MR is consistent throughout, the causality can be firmly established; otherwise, the observed association might be alternatively explained by biases or residual confounders.

## Results

### Cohort evidence

Among the 6,786 participants, most were female (56.4%), with a mean age of 66.3±9.8 years old. People in the high education group were likely to be male and have higher occupation classes and childhood SEPs. Also, they were likely to be non-smokers and had increased physical activity levels and slightly lower BMIs than those in other groups (P-value<0.001). In contrast, those in the low education group tended to be separated, divorced, or widowed and have elevated depressive symptoms and a history of cardiovascular diseases (P-value<0.001, Table 1).

After a median follow-up of 10.4 years, 598 out of the 6,786 participants reported diabetes (10.1 [95%CI 9.3-10.9] per 1,000 person-years). A Kaplan-Meier plot had shown that low education was associated with a significantly higher T2DM incidence (log-rank P-value<0.001, Figure S3). Moreover, we observed a gradient inverse association between the education levels and the risk of T2DM: The hazard ratios (95%CI) of T2DM in the middle and low education groups were 1.20 (0.97-1.49) and 1.58 (1.28-1.96), respectively, compared to the high education group in age, sex, and childhood SEP adjusted models (Table 2). The significance remained after individually adjusting for health behaviours, psychosocial resources, and occupational classes, but the association became null after simultaneous adjustment. Admittedly, sex, age group, BMI, and smoking status did not significantly modify the associations (Figure S4). Furthermore, the observed inverse associations were consistent across sensitivity analyses (Table S4).

**Table 2.** The association between education levels and the incidence of type 2 diabetes mellitus (n=6,786) and the trajectory of HbA1c levels (n=5,158)

| Outcomes | High education | Middle education | Low education |
| --- | --- | --- | --- |
| **Hazard ratios of incident T2DM (95%CI)** |  |  |  |
| Model 1 | 1.00 (ref) | 1.22 (0.99 to 1.51) | **1.71 (1.41 to 2.09)** |
| *Confounder-adjusting models* |  |  |  |
| Model 2 | 1.00 (ref) | **1.28 (1.03 to 1.58)** | **1.78 (1.45 to 2.18)** |
| Model 3<sup>§</sup> | 1.00 (ref) | 1.20 (0.97 to 1.49) | **1.58 (1.28 to 1.96)** |
| *Confounder- and mediator-adjusting models* |  |  |  |
| Model 4 | 1.00 (ref) | 1.10 (0.89 to 1.37) | 1.24 (0.99 to 1.54) |
| Model 5 | 1.00 (ref) | 1.19 (0.96 to 1.48) | **1.54 (1.24 to 1.91)** |
| Model 6 | 1.00 (ref) | 1.16 (0.92 to 1.46) | **1.45 (1.14 to 1.84)** |
| Model 7 | 1.00 (ref) | 1.08 (0.86 to 1.36) | 1.17 (0.92 to 1.50) |
| **β-coefficients of HbA1c levels (95%CI)<sup>‡</sup>** |  |  |  |
| Model 1 | 0.00 (ref) | 0.0263<br>(-0.0048 to 0.0575) | **0.1088**<br>**(0.0772 to 0.1404)** |
| *Confounder-adjusting models* |  |  |  |
| Model 2 | 0.00 (ref) | 0.0231<br>(-0.0081 to 0.0544) | **0.0869**<br>**(0.0543 to 0.1194)** |
| Model 3<sup>§</sup> | 0.00 (ref) | 0.0217<br>(-0.0100 to 0.0533) | **0.0833**<br>**(0.0492 to 0.1174)** |
| *Confounder- and mediator-adjusting models* |  |  |  |
| Model 4 | 0.00 (ref) | 0.0028<br>(-0.0283 to 0.0338) | **0.0396**<br>**(0.0055 to 0.0737)** |
| Model 5 | 0.00 (ref) | 0.0210<br>(-0.0107 to 0.0527) | **0.0815**<br>**(0.0474 to 0.1157)** |
| Model 6 | 0.00 (ref) | 0.0058<br>(-0.0279 to 0.0394) | **0.0554**<br>**(0.0173 to 0.0936)** |
| Model 7 | 0.00 (ref) | -0.0074<br>(-0.0403 to 0.0255) | 0.0230<br>(-0.0148 to 0.0607) |

**Notes:** <sup>§</sup>Represent the main results. <sup>‡</sup>Random-intercept and-slope model with unstructured covariance. Embolden figures to represent statistical significance. **Abbreviations:** HbA1c; Glycated haemoglobin, T2DM; Type-2 diabetes mellitus. **Model 1:** Unadjusted models, **Model 2:** Age and sex-adjusted models, **Model 3:** Model 2 + childhood SEP adjusted models, **Model 4:** Model 3 + health behaviours adjusted models, **Model 5:** Model 4 + psychosocial resources adjusted models, **Model 6:** Model 3 + occupational class adjusted models, **Model 7:** Model 4 + Model 5 + Model 6 adjusted models

Regarding HbA1c levels (Table 2), we noticed that people in a low-education group had slightly higher HbA1c levels than those in a high-education group (β=0.0833, 95%CI 0.0492-0.1174) after controlling for age, sex, and childhood SEP. Additionally, the results were robust after excluding diabetes participants (Table S5). The trajectory of HbA1c levels in each educational group is illustrated in Figure S5.

### Genetic evidence

From 1,271 schooling-associated SNPs, 210 and 184 SNPs were selected and harmonised with T2DM and HbA1c levels, respectively (Figure S6). These can respectively explain 1.6% (F-statistic=88.18) and 1.4% (F-statistic=87.81) of the variability in schooling years.

Although an inverse association between years of schooling and the risk of T2DM was initially observed in the IVW model (Table 3), the results were not robust across sensitivity analyses. In the IVW model, the odds of having T2DM decreased as schooling years increased: OR 0.75 (95%CI 0.57-0.99). The results were consistent with WM: OR 0.64 (95%CI 0.43-0.95) but not with MR Egger (OR 0.81 [95%CI 0.24-2.69]) nor weighted mode MR (OR 0.34 [95%CI 0.11-1.02]). We found no apparent evidence of heterogeneity on T2DM outcome (I^2^=12%, P-value=0.09). Nevertheless, applying Steiger filtering attenuated most results towards the null. Additionally, a scatter plot between SNPs-education and SNPs-T2DM did not show any apparent pattern of the association (Figure S7). We also found a similar way of the associations in HbA1c outcome (Table 3 and Figure S8).

**Table 3.**
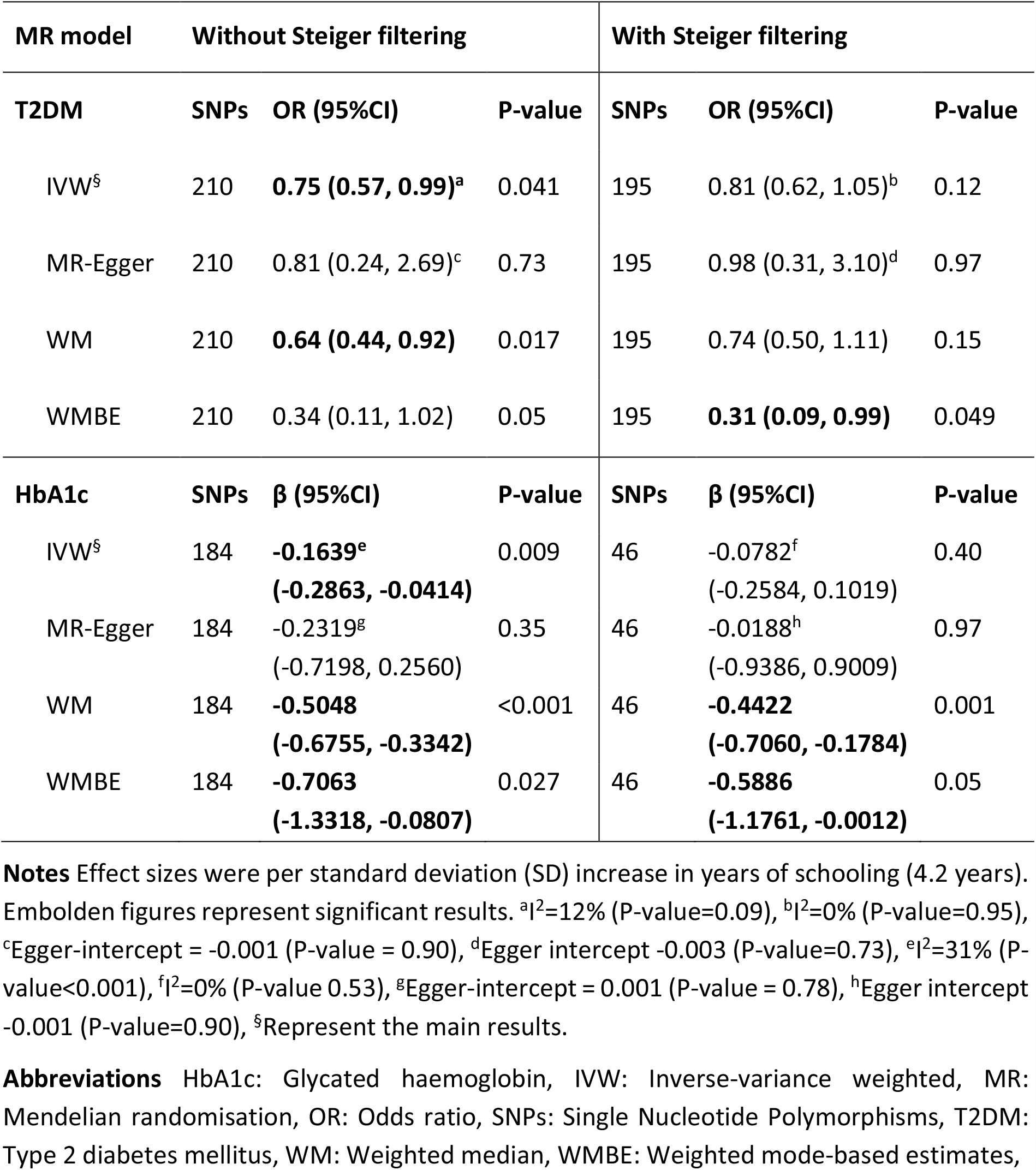
The association between years of schooling, risk of T2DM, and HbA1c levels

Additionally, our positive control showed consistent findings with established evidence, indicating the validity of instruments and processes used in our MR analyses (Table S8).

### Triangulation of evidence

Importantly, when we triangulated pieces of evidence (Figure 1), we found inconsistent results between observational study and MR. While cohort findings suggested inverse associations between education level and the risk of T2DM and HbA1c levels, MR findings suggested null associations.

**Figure 1.**
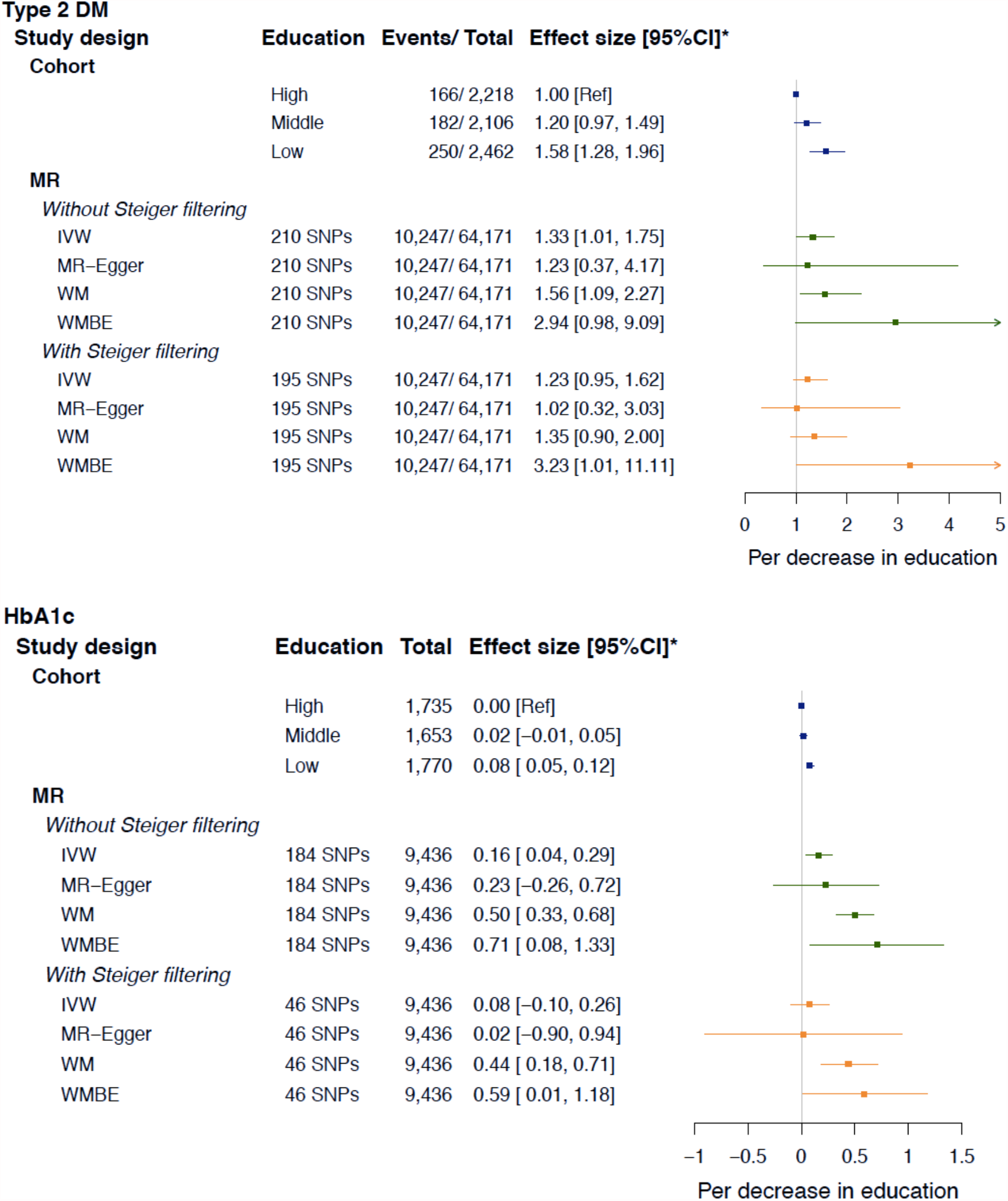
Triangulation of observational and genetic evidence on the association between educational levels and the risk of type 2 diabetes mellitus and HbA1c levels **Notes:** *Effect sizes are hazard ratio (adjusted for age, sex, and childhood SEP) for prospective cohort design and odds ratio for Mendelian randomisation. Effect sizes from MR findings were transformed from the original values to reflect per SD decrease in years of schooling. **Abbreviations**: IVW; Inverse variance weighted, WM; Weighted median, WMBE; Weighted mode-based estimate

## Discussion

### Summary of key findings

In the cohort study, we observed that low education was associated with an increased risk of T2DM, possibly owing to inadequate psychosocial resources, unhealthy behaviours, and a lower occupational class. Moreover, an observed inversed association was the same for HbA1c levels, regardless of T2DM status. Nonetheless, findings from MR did not support a causal association between education and the risk of T2DM and HbA1c levels. Further, they indicated that significant MR results were dominated by SNPs directly associated with the outcome and, therefore, not a good proxy of education.

### Comparing with previous studies

Our observational findings are concordant with previous works indicating that education was inversely associated with incident T2DM, and there is no direct pathway linked to T2DM. However, in contrast to the previous ELSA report,[5] we did not observe different sex-specific associations. This might be because we followed the participants for a more extended period and used HbA1c as an additional criterion to define T2DM. So we could identify more T2DM events in both sexes and gain better statistical power to detect slight differences. Moreover, previous work also used antidiabetic medication data to ascertain diabetes, whereas, in this study, we used only self-reporting diagnosis and HbA1c levels. Nevertheless, when we restricted the analysis to wave 4, we found a trend of the association that was similar to the previous ELSA study, where the association is more substantial in females than in males (results not shown).[5] Also, our results were coherent with previous observational studies.[6–8]

In terms of genetic evidence, an MR study by Hagenaars *et al*. suggested no causal link between educational attainment and the risk of T2DM.[13] However, the study used only 9 SNPs, and the null findings might be due to statistical underpowering. In contrast to ours, two recent MR studies have shown a causal association between education and the risk of T2DM.[11,12] It should be noted that, before applying Steiger filtering, we produced relatively similar (but slightly weaker) results as those works. However, after applying Steiger filtering, almost all MR findings became null. Thus, we cannot exclude the potential direct effect of genetic instruments used in their analyses. In addition, underpowering is unlikely to be a case for T2DM outcome in our MR study (Table S7).

Interestingly, our null findings on HbA1c were consistent with the most recent work by Liang *et al*. despite the contradiction in T2DM outcomes.[14] The discrepancy in results might be due to different methods used for weighting SNPs in the IVW model. Rather than using an additive model, we used a multiplicative one instead, as recommended.[30] The former may upweight outlier SNPs and consequently erroneously strengthen the association, which can be noticed by the similarity of scatter plots of SNPs exposure and SNPs outcome between our work and the previous one.

### Strengths and limitations

To our best knowledge, this is the first report that triangulated cohort and genetic evidence on education and T2DM and HbA1c. However, there are some caveats worth noticing. First, our outcome derived from self-reporting diabetes, which cannot differentiate between T1DM and T2DM, and might be prone to misclassification bias. However, since incident diabetes cases in our study were identified in participants aged ≥50 years, most events were clinically assumed to be T2DM.[10] Additionally, it was shown that self-report diabetes had a very high specificity (99.7%) but low sensitivity (66%).[37] Thus, we used HbA1c as an additional criterion to define the outcome to improve false-negative cases. Second, we cannot exclude the possibility that factors treated as mediators in our analysis can also be confounders since we did not have the exact temporal sequence of each variable. For instance, some participants might already be obese or active smokers before their graduation, and these risks were carried over until age 50 when they participated in ELSA. Lastly, the generalisability of our findings is limited to European ancestry populations.

### Implications of findings

The validity of SNPs used in the MR analysis should be a significant concern for both readers and researchers when interpreting and implementing findings from MR studies. Meanwhile, results from observational research alone prone to being misleading due to biases and residual confounders. Hence, we encouraged using the triangulation approach to gain more confidence in the causality inferences. Also, future works on different ethnicities might warrant generalisability. According to our findings, targeting education might not directly decrease the incidence of T2DM. However, education is a key to improve psychosocial resources, healthy behaviours, and occupation, which might delay the occurrence of T2DM and have a positive impact on health in the long run.[3] Therefore, improving education should still be encouraged, although its causality to T2DM might not exist.

In summary, education did not directly affect T2DM and HbA1c levels. Inadequate psychosocial resources, low occupational class, and unhealthy behaviours could explain the observed inverse associations. Moreover, our triangulation of evidence did not support a causal role of education in the risk of T2DM and HbA1c levels.

## Data Availability

ELSA data were made available through the UK Data Archive. Genetic data used in this research are also publicly available on a public domain.

https://www.ukdataservice.ac.uk/

https://www.mrbase.org/

## Acknowledgements

This research project was supported by the Thailand Science Research and Innovation Fund and the University of Phayao (Grant No. FF64-UoE039). However, the funding body did not involve the design, analysis, and interpretation of this study. The English Longitudinal Study of Ageing (ELSA) is supported by the National Institute on Ageing (grant numbers: 2RO1AG7644 and 2RO1AG017644–01A1) and a consortium of the UK government departments co-ordinated by the Office for National Statistics. Additionally, we would like to thank researchers from the MR-Base Collaboration who made the IEU GWAS database publicly available.

## Declarations

### Availability of data and material

ELSA data were made available through the UK Data Archive (https://www.ukdataservice.ac.uk/). Genetic data used in this research are publicly available from https://www.mrbase.org/.

### Code availability

In this study, all analyses were performed using STATA version 16MR (StataCorp, LLC) package “stcox”, “mixed”, and “mrrobust”. We also used R version 3.6 package “TwoSampleMR” for the genetic instrument extraction and harmonisation. Additional R script and STATA do-file for the analyses were available upon request.

### Disclaimers

The original data creators, depositors or copyright holders, the funders of the Data Collections, and the UK Data Service/UK Data Archive bear no responsibility for analysing or interpreting this study.

### Authors’ contributions

NN conceived the study aims and design and obtained access to ELSA data. NN, JS, and AA contributed to the literature reviewing, data cleaning, data analyses, interpretation of the findings. NN and SB developed the initial and subsequent manuscripts. PC and PD critically revised the initial manuscript, and all authors participated in further revisions. The final manuscript was read and approved by all authors before submission.

### Ethics approval

The English Longitudinal Study of Ageing has been approved by the National Research Ethics Service (London Multicentre Research Ethics Committee (MREC/01/2/91)). For the MR study, specific ethical approval has been obtained individually in the original genome-wide association studies (GWAS).

### Consent to participate

Not applicable (specific consent was obtained in the original studies)

### Consent for publication

Not applicable

