## Supplementary materials for "Educational level as a cause of type 2 diabetes mellitus: Caution from triangulation of observational and genetic evidence"

***Corresponding author**

Dr Nat Na-Ek

Department of Pharmaceutical Care, School of Pharmaceutical Sciences, University of Phayao, Phayao, Thailand,

**Table S8** MR analyses of education and the risk of Alzheimer's disease (positive control) ... 19

**Table S1** STROBE Statement for cohort studies

| | Item No | Recommendation | Page No |
|---|---|---|---|
| **Title and abstract** | 1 | (*a*) Indicate the study's design with a commonly used term in the title or the abstract<br>(*b*) Provide in the abstract an informative and balanced summary of what was done and what was found | 1-2 |
| **Introduction** | | | |
| Background/ rationale | 2 | Explain the scientific background and rationale for the investigation being reported | 3 |
| Objectives | 3 | State specific objectives, including any prespecified hypotheses | 3, 6 |
| **Methods** | | | |
| Study design | 4 | Present key elements of study design early in the paper | 3 |
| Setting | 5 | Describe the setting, locations, and relevant dates, including periods of recruitment, exposure, follow-up, and data collection | 3-4 |
| Participants | 6 | (*a*) Give the eligibility criteria, and the sources and methods of selection of participants. Describe methods of follow-up<br>(*b*) For matched studies, give matching criteria and number of exposed and unexposed | *(a)* 3-4<br>*(b)* NR |
| Variables | 7 | Clearly define all outcomes, exposures, predictors, potential confounders, and effect modifiers. Give diagnostic criteria, if applicable | 4 |
| Data sources/ measurement | 8* | For each variable of interest, give sources of data and details of methods of assessment (measurement). Describe comparability of assessment methods if there is more than one group | 4 |
| Bias | 9 | Describe any efforts to address potential sources of bias | 4-5 |
| Study size | 10 | Explain how the study size was arrived at | NR[$] |
| Quantitative variables | 11 | Explain how quantitative variables were handled in the analyses. If applicable, describe which groupings were chosen and why | 5 |
| Statistical methods | 12 | (*a*) Describe all statistical methods, including those used to control for confounding<br>(*b*) Describe any methods used to examine subgroups and interactions<br>(*c*) Explain how missing data were addressed<br>(*d*) If applicable, explain how loss to follow-up was addressed<br>(*e*) Describe any sensitivity analyses | *(a)* - *(e)* 5 |
| **Results** | | | |
| Participants | 13* | (a) Report numbers of individuals at each stage of study—eg numbers potentially eligible, examined for eligibility, confirmed eligible, included in the study, completing follow-up, and analysed<br>(b) Give reasons for non-participation at each stage<br>(c) Consider use of a flow diagram | (*a*) – (*c*) Figure S1 |
| Descriptive data | 14* | (a) Give characteristics of study participants (eg demographic, clinical, social) and information on exposures and potential confounders<br>(b) Indicate number of participants with missing data for each variable of interest<br>(c) Summarise follow-up time (eg, average and total amount) | (*a*) - (*c*) 7, Table 1 |
| Outcome data | 15* | Report numbers of outcome events or summary measures over time | 7, Fig S3 |

| | | | |
|---|---|---|---|
| Main results | 16 | (*a*) Give unadjusted estimates and, if applicable, confounder-adjusted estimates and their precision (eg, 95% confidence interval). Make clear which confounders were adjusted for and why they were included<br>(*b*) Report category boundaries when continuous variables were categorized<br>(*c*) If relevant, consider translating estimates of relative risk into absolute risk for a meaningful time period | (*a*) 7, Table 2, Figure S2<br><br>(*b*)-(*c*) NR |
| Other analyses | 17 | Report other analyses done—eg analyses of subgroups and interactions, and sensitivity analyses | Figure S4, Table S3-S5 |
| **Discussion** | | | |
| Key results | 18 | Summarise key results with reference to study objectives | 8 |
| Limitations | 19 | Discuss limitations of the study, taking into account sources of potential bias or imprecision. Discuss both direction and magnitude of any potential bias | 9 |
| Interpretation | 20 | Give a cautious overall interpretation of results considering objectives, limitations, multiplicity of analyses, results from similar studies, and other relevant evidence | 8-9 |
| Generalisability | 21 | Discuss the generalisability (external validity) of the study results | 9 |
| **Other information** | | | |
| Funding | 22 | Give the source of funding and the role of the funders for the present study and, if applicable, for the original study on which the present article is based | 10 |

*Give information separately for exposed and unexposed groups.
$All eligible participants were included in the analyses and the results were statistically significant.
Abbreviation: NR; Not relevant

**Note:** An Explanation and Elaboration article discusses each checklist item and gives methodological background and published examples of transparent reporting. The STROBE checklist is best used in conjunction with this article (freely available on the Web sites of PLoS Medicine at http://www.plosmedicine.org/, Annals of Internal Medicine at http://www.annals.org/, and Epidemiology at http://www.epidem.com/). Information on the STROBE Initiative is available at http://www.strobe-statement.org.

**Table S2** STROBE-MR Statement for Mendelian randomisation studies

| | Item No | Recommendation | Page No |
|---|---|---|---|
| **Title and abstract** | 1 | Indicate Mendelian randomization as the study's design in the title and/or the abstract | 1-2 |
| **Introduction** | | | |
| Background/rationale | 2 | Explain the scientific background and rationale for the reported study. Is causality between exposure and outcome plausible? Justify why MR is a helpful method to address the study question. | 3 |
| Objectives | 3 | State specific objectives clearly, including prespecified causal hypotheses (if any). | 3, 6 |
| **Methods** | | | |
| Study design | 4 | Present key elements of study design early in the paper. Consider including a table listing sources of data for all phases of the study. For each data source contributing to the analysis, describe the following:<br>(*a*) Describe the study design and the underlying population from which it was drawn. Describe also the setting, locations, and relevant dates, including periods of recruitment, exposure, follow-up, and data collection, if available.<br>(*b*) Give the eligibility criteria, and the sources and methods of selection of participants.<br>(*c*) Explain how the analyzed sample size was arrived at.<br>(*d*) Describe measurement, quality and selection of genetic variants.<br>(*e*) For each exposure, outcome and other relevant variables, describe methods of assessment and, in the case of diseases, the diagnostic criteria used.<br>(*f*) Provide details of ethics committee approval and participant informed consent, if relevant. | (*a*) - (*b*) 5, Table S6<br>(*c*) Table S7<br>(*d*) 5<br>(*e*) 6<br>(*f*) NR |
| Assumptions | 5 | Explicitly state assumptions for the main analysis (e.g. relevance, exclusion, independence, homogeneity) as well assumptions for any additional or sensitivity analysis. | 6, Suppl page 8 |
| Statistical methods: main analysis | 6 | Describe statistical methods and statistical used.<br>(*a*) Describe how quantitative variables were handled in the analyses (i.e., scale, units, model).<br>(*b*) Describe the process for identifying genetic variants and weights to be included in the analyses (i.e, independence and model). Consider a flow diagram.<br>(*c*) Describe the MR estimator, e.g. two-stage least squares, Wald ratio, and related statistics. Detail the included covariates and, in case of two-sample MR, whether the same covariate set was used for adjustment in the two samples.<br>(*d*) Explain how missing data were addressed.<br>(*e*) If applicable, say how multiple testing was dealt with. | (*a*) 5<br>(*b*) 5-6, Figure S6<br>(*c*) Table S6<br>(*d*) NR<br>(*e*) 6 |
| Assessment of assumptions | 7 | Describe any methods used to assess the assumptions or justify their validity. | 6, Suppl page 8 |
| Sensitivity analyses | 8 | Describe any sensitivity analyses or additional analyses performed. | 6 |
| Software and pre-registration | 9 | (*a*) Name statistical software and package(s), including version and settings used.<br>(*b*) State whether the study protocol and details were pre-registered (as well as when and where). | (*a*) 6<br>(*b*) NR |
| **Results** | | | |

| | Item No | Recommendation | Page No |
|---|---|---|---|
| Descriptive data | 10 | (*a*) Report the numbers of individuals at each stage of included studies and reasons for exclusion. Consider use of a flow-diagram.<br>(*b*) Report summary statistics for phenotypic exposure(s), outcome(s) and other relevant variables (e.g. means, standard deviations, proportions).<br>I If the data sources include meta-analyses of previous studies, provide the number of studies, their reported ancestry, if available, and assessments of heterogeneity across these studies. Consider using a supplementary table for each data source.<br>(*d*) For two-sample Mendelian randomization:<br>i. Provide information on the similarity of the genetic variant-exposure associations between the exposure and outcome samples.<br>ii. Provide information on extent of sample overlap between the exposure and outcome data sources. | (*a*)-(*c*) NR<br>(*d*) Not report |
| Main results | 11 | (a) Report the associations between genetic variant and exposure, and between genetic variant and outcome, preferably on an interpretable scale (e.g. comparing 25$^{th}$ and 75$^{th}$ percentile of allele count or genetic risk score, if individual-level data available). | Table 3 |
| | | (b) Report causal effect estimate between exposure and outcome, and the measures of uncertainty from the MR analysis. Use an intuitive scale, such as odds ratio, or relative risk, per standard deviation difference. | Table 3, Figure 1 |
| | | (c) If relevant, consider translating estimates of relative risk into absolute risk for a meaningful time-period. | NR |
| | | (d) Consider any plots to visualize results (e.g. forest plot, scatterplot of associations between genetic variants and outcome versus between genetic variants and exposure). | Figure 1, Figure S7-S8 |
| Assessment of assumptions | 12 | (a) Assess the validity of the assumptions. | 7-8 |
| | | (b) Report any additional statistics (e.g., assessments of heterogeneity, such as I2, Q statistic). | 7-8, Table 3 |
| Sensitivity and additional analyses | 13 | (a) Use sensitivity analyses to assess the robustness of the main results to violations of the assumptions. | 7-8, Table 3 |
| | | (b) Report results from other sensitivity analyses (e.g., replication study with different dataset, analyses of subgroups, validation of instrument(s), simulations, etc.). | Table S8 |
| | | (c) Report any assessment of direction of causality (e.g., bidirectional MR). | Table 3 |
| | | (d) When relevant, report and compare with estimates from non-MR analyses. | Figure 1 |
| | | (e) Consider any additional plots to visualize results (e.g., leave-one-out analyses). | Not report |
| Discussion | | | |
| Key results | 14 | Summarize key results with reference to study objectives | 8 |
| Limitations | 15 | Discuss limitations of the study, taking into account the validity of the MR assumptions, other sources of potential bias, and imprecision. Discuss both direction and magnitude of any potential bias, and any efforts to address them. | 8-9 |
| Interpretation | 16 | (a) Give a cautious overall interpretation of results considering objectives and limitations. Compare with results from other relevant studies. | 8-9 |
| | | (b) Discuss underlying biological mechanisms that could be modelled by using the genetic variants to assess the relationship between the exposure and the outcome. | NR |

| | Item No | Recommendation | Page No |
|---|---|---|---|
| | | (c) Discuss whether the results have clinical or policy relevance, and whether interventions could have the same size effect. | 9 |
| Generalizability | 17 | Discuss the generalizability of the study results (a) to other populations (i.e., external validity), (b) across other exposure periods/timings, and (c) across other levels of exposure | 9 |
| **Other information** | | | |
| Funding | 18 | Give the source of funding and the role of the funders for the present study and, if applicable, for the original study or studies on which the present article is based. | 10 |
| Data and data sharing | 19 | Present data used to perform all analyses or report where and how the data can be accessed. State whether statistical code is publicly accessible and if so, where. | 10 |
| Conflicts of Interest | 20 | All authors should declare all potential conflicts of interest. | 10 |

**Note**: The above table was created based on Smith et al. [1]

**A conceptual framework to obtain a conclusion from the MR analysis of this study**

Adapted from Burgess *et al.*[2]

- The IVW is the primary method to exclude the conclusion that there is no firm evidence of a causal effect. If the IVW results are significant, additional analyses will help ensure the robustness of significant findings.
- MR Egger is used to check and control for unbalanced horizontal pleiotropy. If the Egger intercept significantly deviated from zero, MR Egger would provide less bias than the IVW method.
- If additional analyses, such as weighted median MR and weighted mode-based MR, showed consistent findings throughout, we would have more confidence in the causal effect. Otherwise, the causality is unlikely.

**Table S3** Factors associated with incident T2DM

| Characteristics | T2DM | | Total | P-value |
|---|---|---|---|---|
| | No | Yes | | |
| N | 6,188 | 598 | 6,786 | |
| Male | 2,654 (42.9%) | 307 (51.3%) | 2,961 (43.6%) | < 0.001 |
| Age (years) | 66.33 ± 0.13 | 65.61 ± 0.35 | 66.27 ± 0.12 | 0.05 |
| Occupation class | | | | |
| High | 1,975 (31.9%) | 168 (28.1%) | 2,143 (31.6%) | < 0.001 |
| Middle | 1,608 (26.0%) | 130 (21.7%) | 1,738 (25.6%) | $P_{trend}$ < 0.001 |
| Low | 2,520 (40.7%) | 293 (49.0%) | 2,813 (41.5%) | |
| Missing | 85 (1.4%) | 7 (1.2%) | 92 (1.4%) | |
| Childhood socioeconomic position | | | | |
| High | 1,945 (31.4%) | 142 (23.7%) | 2,087 (30.7%) | < 0.001 |
| Middle | 2,019 (32.6%) | 197 (32.9%) | 2,216 (32.7%) | $P_{trend}$ < 0.001 |
| Low | 1,981 (32.0%) | 236 (39.5%) | 2,217 (32.7%) | |
| Miscellaneous (included missing) | 243 (3.9%) | 23 (3.8%) | 266 (3.9%) | |
| Marital status | | | | |
| Single | 330 (5.3%) | 23 (3.8%) | 353 (5.2%) | 0.29 |
| Married | 4,084 (66.0%) | 398 (66.6%) | 4,482 (66.0%) | |
| Separated, divorced, widowed | 1,774 (28.7%) | 177 (29.6%) | 1,951 (28.8%) | |
| Having depressive symptoms | 857 (13.8%) | 96 (16.1%) | 953 (14.0%) | 0.14 |
| Missing | 40 (0.6%) | 4 (0.7%) | 44 (0.6%) | |
| Smoking status | | | | |
| Never smoked | 2,310 (37.3%) | 200 (33.4%) | 2,510 (37.0%) | 0.003 |
| Ex-smokers | 2,964 (47.9%) | 276 (46.2%) | 3,240 (47.7%) | $P_{trend}$ = 0.003 |
| Current smokers | 892 (14.4%) | 116 (19.4%) | 1,008 (14.9%) | |
| Missing | 22 (0.4%) | 6 (1.0%) | 28 (0.4%) | |
| Alcohol drinking status | | | | |
| Never/ almost never | 1,626 (26.3%) | 181 (30.3%) | 1,807 (26.6%) | <0.001 |
| 1-2 times/ month | 652 (10.5%) | 90 (15.1%) | 742 (10.9%) | $P_{trend}$ < 0.001 |
| 1-2 times/ week | 1,975 (31.9%) | 193 (32.3%) | 2,168 (31.9%) | |
| Daily/ almost daily | 1,912 (30.9%) | 127 (21.2%) | 2,039 (30.0%) | |
| Missing | 23 (0.4%) | 7 (1.2%) | 30 (0.4%) | |
| Physical activity level | | | | |
| High | 1,275 (20.6%) | 89 (14.9%) | 1,364 (20.1%) | <0.001 |
| Middle | 3,210 (51.9%) | 305 (51.0%) | 3,515 (51.8%) | $P_{trend}$ < 0.001 |
| Low | 1,396 (22.6%) | 178 (29.8%) | 1,574 (23.2%) | |
| Sedentary | 264 (4.3%) | 22 (3.7%) | 286 (4.2%) | |

| Characteristics | T2DM | | Total | P-value |
|---|---|---|---|---|
| | No | Yes | | |
| Missing | 43 (0.7%) | 4 (0.7%) | 47 (0.7%) | |
| Body mass index (kg/m$^2$) | 27.36 ± 0.07 | 30.87 ± 0.25 | 27.67 ± 0.07 | <0.001 |
| Normal | 1,393 (22.5%) | 46 (7.7%) | 1,439 (21.2%) | <0.001 |
| Overweight | 2,144 (34.6%) | 170 (28.4%) | 2,314 (34.1%) | $P_{trend}$ < 0.001 |
| Obesity | 1,173 (19.0%) | 239 (40.0%) | 1,412 (20.8%) | |
| Missing | 1,478 (23.9%) | 143 (23.9%) | 1,621 (23.9%) | |
| Having a history of CVDs | 3,125 (50.5%) | 375 (62.7%) | 3,500 (51.6%) | <0.001 |

**Notes** Figures represent frequency (%) or mean ± SD.

**Table S4** Sensitivity analysis: Complete-case analysis and Bonferroni adjusted confidence interval

| **Model** | **Hazard ratio of incident T2DM (97.5%CI)** | | |
|---|---|---|---|
| | **High education** | **Middle education** | **Low education** |
| **Complete-case analysis (n=4,951)** | | | |
| Model 1 | 1.00 (reference) | 1.17 (0.89 to 1.54) | **1.69 (1.30 to 2.19)** |
| Model 2 | 1.00 (reference) | 1.23 (0.93 to 1.62) | **1.74 (1.32 to 2.28)** |
| Model 3§ | 1.00 (reference) | 1.14 (0.86 to 1.52) | **1.52 (1.14 to 2.01)** |
| Model 4 | 1.00 (reference) | 1.08 (0.81 to 1.43) | 1.23 (0.92 to 1.65) |
| Model 5 | 1.00 (reference) | 1.13 (0.85 to 1.50) | **1.49 (1.12 to 1.98)** |
| Model 6 | 1.00 (reference) | 1.06 (0.79 to 1.43) | 1.32 (0.96 to 1.81) |
| Model 7 | 1.00 (reference) | 1.01 (0.74 to 1.36) | 1.12 (0.81 to 1.54) |
| **Multiply imputed analysis (n=6,786)** | | | |
| Model 1 | 1.00 (reference) | 1.22 (0.96 to 1.55) | **1.71 (1.37 to 2.15)** |
| Model 2 | 1.00 (reference) | **1.28 (1.01 to 1.63)** | **1.78 (1.41 to 2.24)** |
| Model 3§ | 1.00 (reference) | **1.20 (0.94 to 1.54)** | **1.58 (1.24 to 2.02)** |
| Model 4 | 1.00 (reference) | 1.10 (0.86 to 1.42) | 1.24 (0.96 to 1.59) |
| Model 5 | 1.00 (reference) | 1.19 (0.93 to 1.52) | **1.54 (1.21 to 1.97)** |
| Model 6 | 1.00 (reference) | 1.16 (0.89 to 1.50) | **1.45 (1.10 to 1.90)** |
| Model 7 | 1.00 (reference) | 1.08 (0.83 to 1.41) | 1.17 (0.89 to 1.56) |

**Notes** Embolden figures represent statistically significant values. §Represent the main results.

**Model 1**: Unadjusted model, **Model 2**: Age and sex adjusted model, **Model 3**: Model 2 with further adjusting for childhood SEP, **Model 4**: Model 3 with further adjusting for health behaviours, including body mass index, smoking, alcohol drinking, and physical activity, **Model 5**: Model 3 with further adjusting for psychosocial resources, including depressive symptoms and marital status, **Model 6**: Model 3 with further adjusting for occupational class, **Model 7**: Model 4 + Model 5 + Model 6

**Table S5** The association between education levels and the trajectory of HbA1c levels after excluding diabetes patients (n=4,741)

| Model | β-coefficient of HbA1c levels (95%CI) | | |
|---|---|---|---|
| | **High education** | **Middle education** | **Low education** |
| Model 1 | 0.0000 (reference) | 0.0188 (-0.0058 to 0.0434) | **0.0865 (0.0611 to 0.1120)** |
| Model 2 | 0.0000 (reference) | 0.0095 (-0.0150 to 0.0340) | **0.0518 (0.0257 to 0.0779)** |
| Model 3§ | 0.0000 (reference) | 0.0099 (-0.0149 to 0.0348) | **0.0524 (0.0251 to 0.0798)** |
| Model 4 | 0.0000 (reference) | -0.0030 (-0.0276 to 0.0215) | 0.0214 (-0.0061 to 0.0489) |
| Model 5 | 0.0000 (reference) | 0.0098 (-0.0150 to 0.0347) | **0.0515 (0.0241 to 0.0789)** |
| Model 6 | 0.0000 (reference) | 0.0008 (-0.0256 to 0.0272) | **0.0365 (0.0059 to 0.0671)** |
| Model 7 | 0.0000 (reference) | -0.0076 (-0.0337 to 0.0185) | 0.0135 (-0.0169 to 0.0439) |

**Notes** Random-intercept and-slope linear mixed model with unstructured covariance. Embolden figures to represent statistical significance. §Represent the main results.

**Model 1**: Unadjusted model, **Model 2**: Age and sex-adjusted model, **Model 3**: Model 2 with further adjusting for childhood SEP, **Model 4**: Model 3 with further adjusting for health behaviours, including body mass index, smoking, alcohol drinking, and physical activity, **Model 5**: Model 3 with further adjusting for psychosocial resources, including depressive symptoms and marital status, **Model 6**: Model 3 with further adjusting for occupational class, **Model 7**: Model 4 + Model 5 + Model 6

8,780 Core members in ELSA wave 2

206 Participants were excluded due to being non-white.

1,788 Participants were further excluded due to:

- Missing data on main exposure (i.e., education) at baseline (n=6)
- Having history of DM or high blood sugar at baseline (n=1,782)

6,786 Participants included in the final analysis

- 6,786 Participants included in the analysis of the primary outcome (i.e., incident diabetes)
- 5,158 Participants who had HbA1c measured at least once during follow-up included in the analysis of secondary outcome (i.e., HbA1c levels)

**Figure S1** Study populations' flow diagram

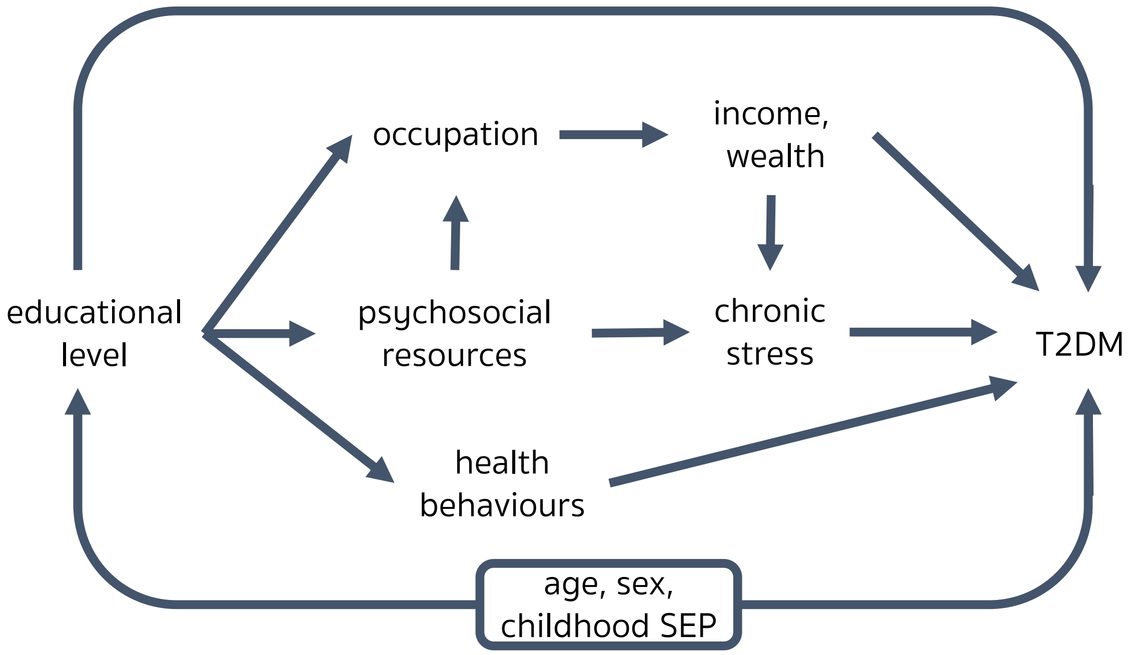

**Figure S2** Directed acyclic graphs (DAGs)

**Notes**: DAGs were adapted from Hamad *et al*.[3] and Liang *et al*.[4] **Abbreviations**: T2DM; Type 2 diabetes mellitus, SEP; Socioeconomic position

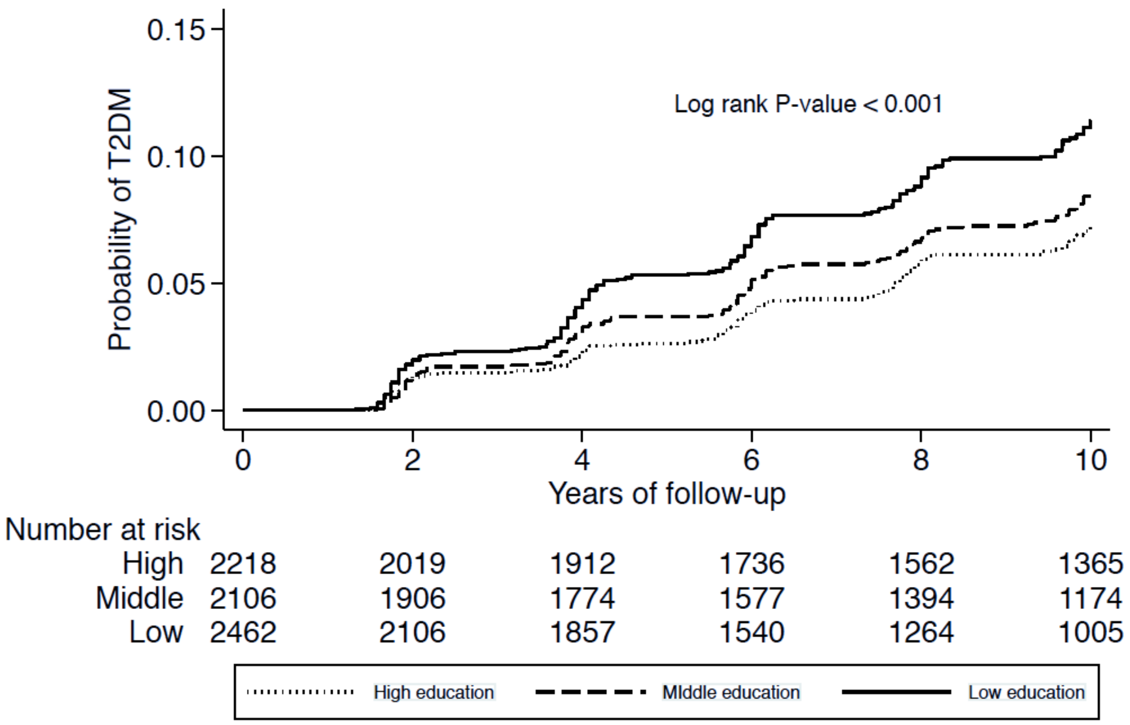

**Figure S3** Kaplan-Meier curve of T2DM according to educational levels

| Characteristics | Subgroups | Education | HR [95%CI] | P–value for interaction |
|---|---|---|---|---|
| **Sex** | Male | High | 1.00 [Ref] | 0.67 |
| | | Middle | 1.14 [0.85, 1.52] | |
| | | Low | 1.60 [1.20, 2.12] | |
| | Female | High | 1.00 [Ref] | |
| | | Middle | 1.32 [0.94, 1.84] | |
| | | Low | 1.59 [1.14, 2.22] | |
| **Age** | < 75 | High | 1.00 [Ref] | 0.14 |
| | | Middle | 1.28 [1.01, 1.61] | |
| | | Low | 1.72 [1.36, 2.17] | |
| | 75+ | High | 1.00 [Ref] | |
| | | Middle | 0.77 [0.42, 1.41] | |
| | | Low | 0.92 [0.54, 1.57] | |
| **Body mass index** | Normal | High | 1.00 [Ref] | 0.60 |
| | | Middle | 1.41 [0.66, 3.04] | |
| | | Low | 1.46 [0.65, 3.27] | |
| | Overweight | High | 1.00 [Ref] | |
| | | Middle | 0.86 [0.59, 1.26] | |
| | | Low | 0.94 [0.64, 1.39] | |
| | Obesity | High | 1.00 [Ref] | |
| | | Middle | 1.44 [1.01, 2.05] | |
| | | Low | 1.80 [1.28, 2.55] | |
| **Smoking status** | Never | High | 1.00 [Ref] | 0.58 |
| | | Middle | 0.99 [0.69, 1.42] | |
| | | Low | 1.35 [0.94, 1.95] | |
| | Ex– | High | 1.00 [Ref] | |
| | | Middle | 1.34 [0.98, 1.82] | |
| | | Low | 1.52 [1.11, 2.10] | |
| | Current | High | 1.00 [Ref] | |
| | | Middle | 1.35 [0.76, 2.40] | |
| | | Low | 1.92 [1.12, 3.29] | |

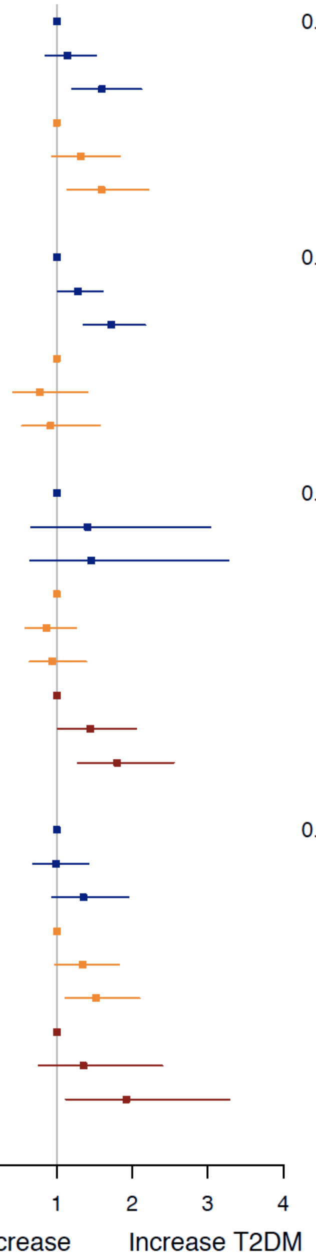

**Figure S4** Subgroup analysis of observational evidence on educational levels and the risk of T2DM

(A) Main analysis (n=5,158)

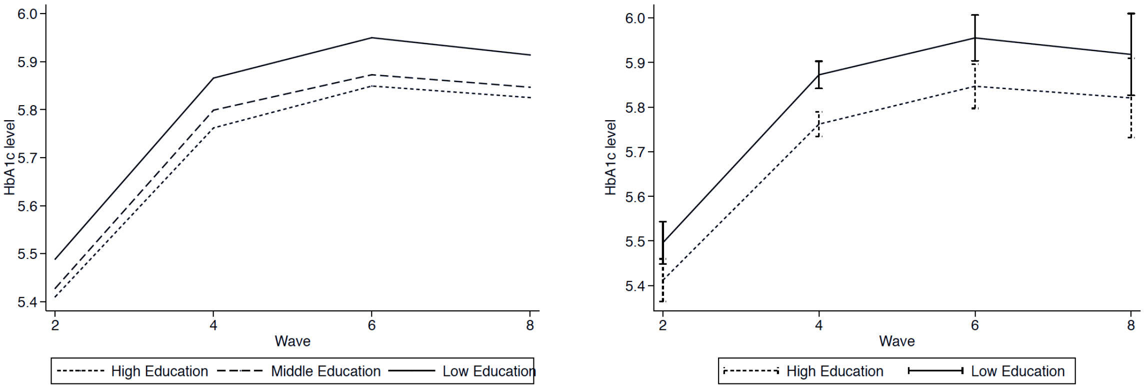

(B) After excluding diabetic patients (n=4,741)

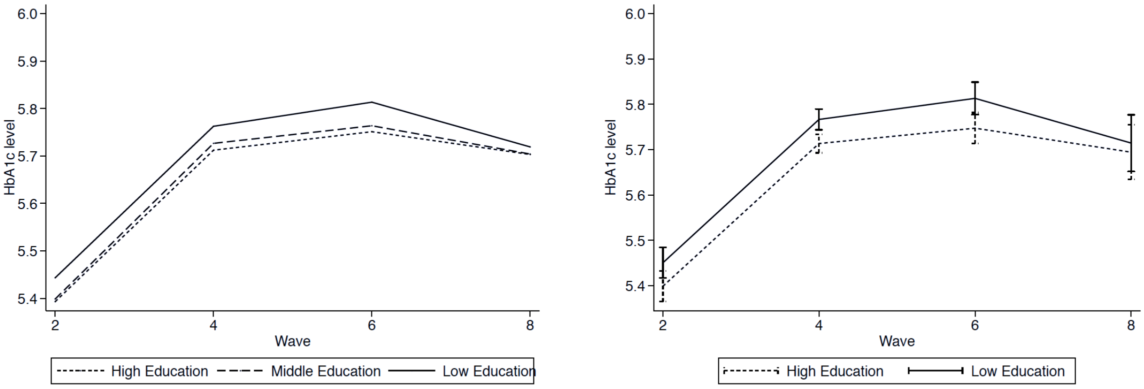

**Figure S5** Trajectory of HbA1c levels across eight waves of follow-up according to educational levels

**Notes**: Main analysis (A), and after excluding individuals with reported diabetes (B). Models were derived from a random intercept and random slope linear mixed model adjusting for age sex and childhood SEP at baseline (ELSA wave 2) with an unstructured covariance

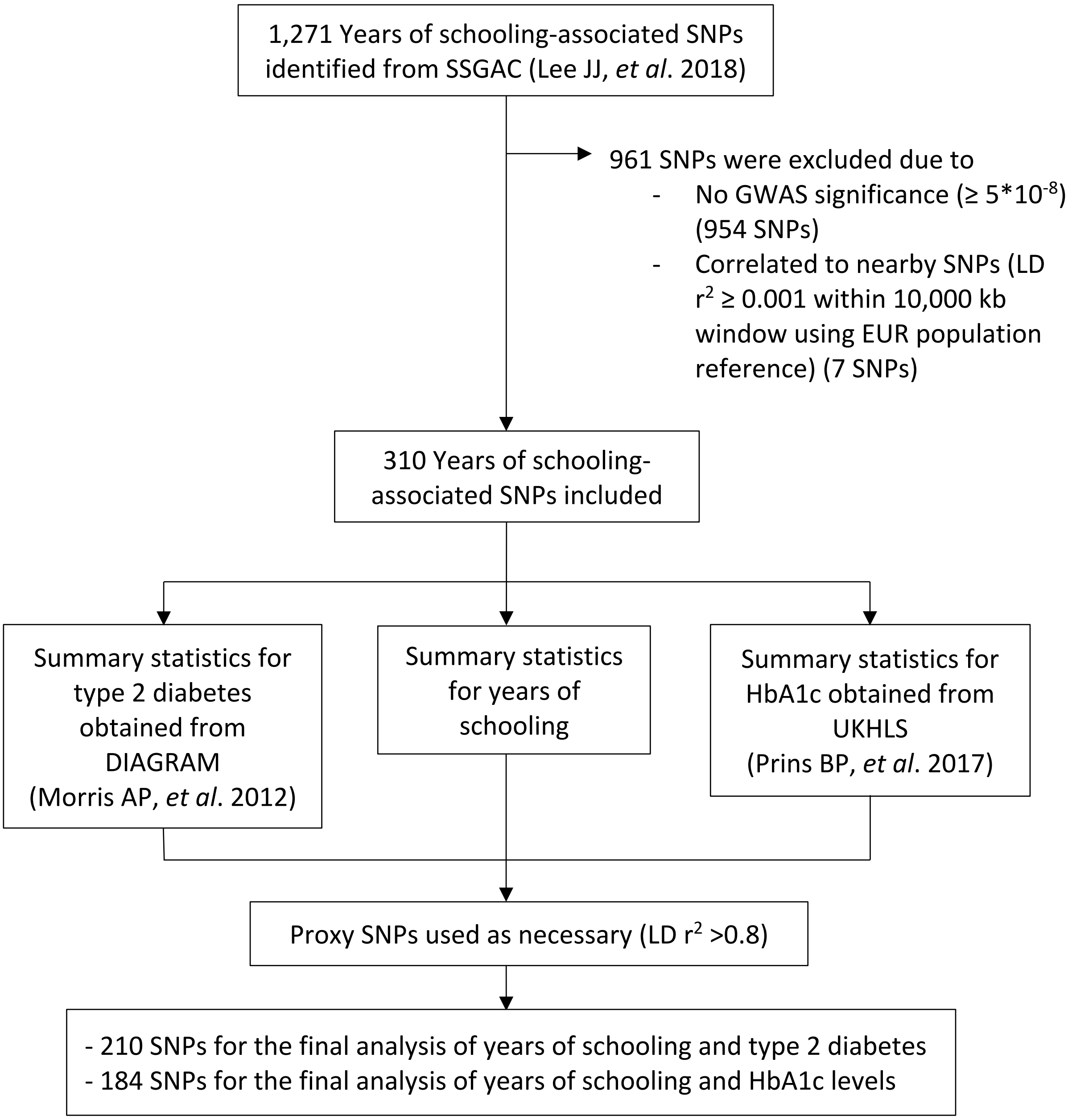

**Figure S6** SNPs' flow diagram

**Table S6** Details of genetic consortia used in the MR analyses

| Consortium | Trait/Disease | No. of participants | Participants' descent |
|---|---|---|---|
| SSGAC | Years of schooling (Per SD [4.2 years] increase) | 1,131,881 | European ancestry |
| DIAGRAM | Type 2 diabetes | Cases: 34,840<br>Controls: 114,981 | European ancestry |
| UKHLS | Non-fasting HbA1c (%, NGSP) | 9,961 | European ancestry |

**Notes:** All consortia were publicly available at https://www.mrbase.org/.

**Table S7** Power calculation of the MR analyses

| Trait | T2DM (cases/controls = 34,840/114,981) | | | HbA1c (sample size = 9,961) | | |
|---|---|---|---|---|---|---|
| | **No. SNPs** | **$r^2$ exposure** | **OR** | **No. SNPs** | **$r^2$ exposure** | **β** |
| Without Steiger filtering | 210 | 0.0161 | 0.872 | 184 | 0.0119 | -0.5192 |
| With Steiger filtering | 195 | 0.0129 | 0.859 | 46 | 0.0038 | -0.9113 |

**Notes**: OR and β represent the minimum effect size detected in our study with sufficient (≥80%) power. **Abbreviations**: HbA1c; Glycated haemoglobin, OR; Odds ratio, SNPs; Single-nucleotide polymorphisms, T2DM; Type 2 diabetes mellitus

**Table S8** MR analyses of education and the risk of Alzheimer's disease (positive control)

| MR models | Our analyses† | | Larsson *et al.*[5]‡ |
|---|---|---|---|
| | No Steiger filtering | Steiger filtering | |
| Number of SNPs | 258 | 237 | 152 |
| IVW | 0.67 (0.57, 0.80) | 0.69 (0.58, 0.82) | 0.89 (0.84, 0.93) |
| MR-Egger | 0.48 (0.24, 0.97) | 0.42 (0.21, 0.83) | 0.72 (0.56, 0.94) |
| Weighted median | 0.61 (0.48, 0.79) | 0.64 (0.50, 0.82) | 0.91 (0.84, 0.98) |
| Mode-based estimate | 0.35 (0.16, 0.74) | 0.32 (0.15, 0.69) | 0.91 (0.84, 0.98)§ |

**Notes**: Estimates (Odds ratio [95%CI]) are per 1 SD increased years of education (†) or per year of schooling completed(‡). **Abbreviations**: IVW; Inverse-variance weighted, MR; Mendelian randomisation, SNPs; Single-nucleotide polymorphisms, §Penalised weighted median

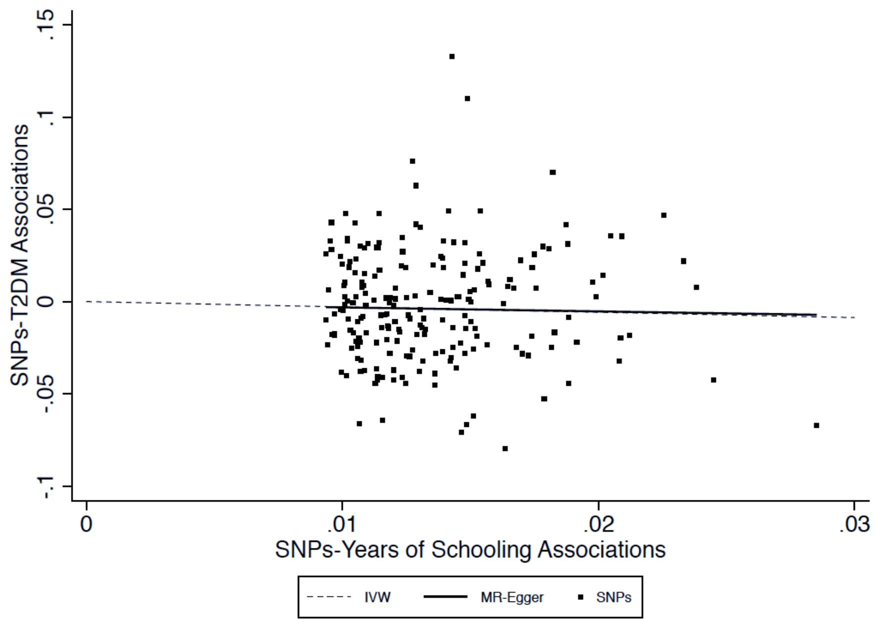

**Figure S7** Scatter plot of SNPs-years of schooling and SNPs-risk of T2DM

**Abbreviations** IVW: Inverse-variance weighted, SNPs: Single Nucleotide Polymorphisms, T2DM: Type 2 diabetes mellitus

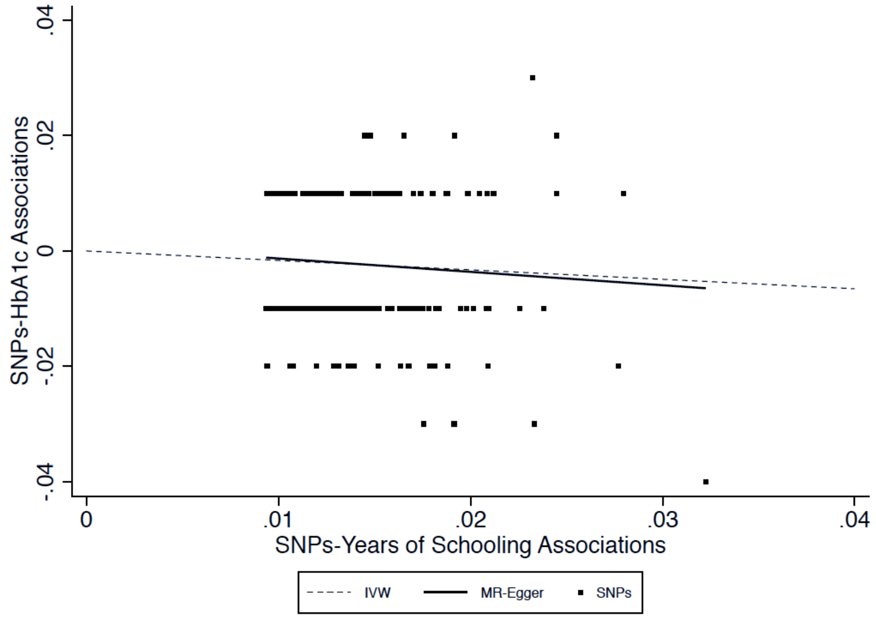

**Figure S8** Scatter plot of SNPs-years of schooling and SNPs-HbA1c levels

**Abbreviations** IVW: Inverse-variance weighted, SNPs: Single Nucleotide Polymorphisms, HbA1c: Glycated haemoglobin
